# Association of chronic heart failure and its comorbidities with loss of actuarially predicted life expectancy: a prospective cohort study

**DOI:** 10.1101/2020.07.02.20145011

**Authors:** Michael Drozd, Samuel D Relton, Andrew MN Walker, Thomas Slater, John Gierula, Maria F Paton, Judith Lowry, Sam Straw, Aaron Koshy, Melanie McGinlay, Alexander D Simms, V Kate Gatenby, Robert J Sapsford, Klaus K Witte, Mark T Kearney, Richard M Cubbon

**Affiliations:** Leeds Institute of Cardiovascular and Metabolic Medicine, The University of Leeds, Clarendon Way, Leeds, LS2 9JT, United Kingdom; Leeds Institute of Health Sciences, The University of Leeds, Clarendon Way, Leeds, LS2 9JT, United Kingdom; Department of Cardiology, Leeds General Infirmary, Leeds Teaching Hospitals NHS Trust, Great George Street, Leeds, LS1 3EX, United Kingdom

**Keywords:** Heart failure, survival, mortality, comorbidity, life expectancy

## Abstract

**Background:** Estimating survival can aid care planning, but the use of absolute survival projections can be challenging for patients and clinicians to contextualize. We aimed to define how heart failure and its major comorbidities contribute to loss of actuarially predicted life expectancy.

**Methods:** We conducted an observational cohort study of 1794 adults with stable chronic heart failure and reduced left ventricular ejection fraction, recruited from cardiology outpatient departments of 4 United Kingdom (UK) hospitals. Data from an 11-year maximum (5-year median) follow-up period (999 deaths) was used to define how heart failure and its major comorbidities impact upon survival, relative to an age-sex matched control UK population, using a relative survival framework.

**Results:** After 10 years, mortality in the reference control population was 29%. In people with heart failure, this increased by an additional 37% (95% confidence interval 34-40%), equating to an additional 2.2-years of lost life, or a 2.4-fold (2.2-2.5) excess loss of life. This excess was greater in men than women (2.4 years [2.2-2.7] versus 1.6 years [1.2-2.0]; p<0.001). In patients without major comorbidity, men still experienced excess loss of life, whilst women experienced less and were non-significantly different from the reference population (1 year [0.6-1.5] versus 0.4 years [-0.3-1]; p<0.001). Accrual of comorbidity was associated with substantial increases in excess loss of life, particularly for chronic kidney and lung disease.

**Conclusions:** Comorbidity accounts for the majority of lost life expectancy in people with heart failure. Women, but not men, without comorbidity experience survival close to reference controls.

## Background

Chronic heart failure (CHF) is a common late phase in the natural history of many cardiovascular diseases, affecting millions of people globally, and remains associated with an appreciable mortality rate (1). In spite of declining age-sex adjusted incidence rates, the prevalence of heart failure continues to increase (2), reflecting improving survival rates and an ageing population. Hence, people with heart failure are increasingly old and have a rising burden of major comorbidity (2). These trends pose challenges for the estimation and communication of prognosis, with important implications for patients and clinicians aiming to make well-informed decisions. For example, established prognostication tools may be less reliable at predicting remaining life expectancy in people over 80 (3), and do not convey the substantial risk of death in similarly aged individuals without heart failure. Moreover, prognostic estimates do not describe the relative contribution of heart failure versus associated comorbidities, which may be important in defining therapeutic priorities in the growing population with multimorbidity. Indeed, non-cardiovascular causes of death are increasingly common in people with heart failure, especially with advancing age (4,5). Furthermore, prior research has shown substantial discordance between patient-predicted and prognostic model predicted survival, illustrating the need to better communicate this important and sensitive topic (6). These issues suggest that alternate approaches to considering and communicating prognosis may be helpful for health professionals and people with heart failure. Therefore, we set out to describe the survival of people with heart failure *relative* to an age-sex matched control population and then define how comorbid disease contributes to the observed loss of survival.

## Methods

As described in our earlier publications (4), we conducted a prospective cohort study with the pre-defined aim of identifying prognostic markers in patients with CHF and reduced left ventricular ejection fraction (LVEF), receiving contemporary evidence-based therapy. Inclusion in the study required the presence of stable signs and symptoms of CHF for at least 3 months, age ≥18 years, and LVEF ≤45% on transthoracic echocardiography. Between June 2006 and December 2014, consecutive patients attending specialist cardiology clinics (secondary and tertiary referral) in four United Kingdom (UK) hospitals were approached, and 1794 patients provided written informed consent. The Leeds West Research Ethics Committee gave ethical approval and the investigation conforms to the principles outlined in the *Declaration of Helsinki*.

Details of comorbid diabetes and chronic obstructive pulmonary disease (COPD) were collected at recruitment, and symptomatic status was defined using the New York Heart Association (NYHA) classification (4). Venous blood was collected at study recruitment for assessment of renal function in the local hospital chemical pathology laboratories. Estimated glomerular filtration rate (eGFR) was calculated using the Modification of Diet in Renal Disease method, with chronic kidney disease (CKD) stage 4 or worse being defined as eGFR<30 ml/minute/1.73m^2^ (7). Two-dimensional echocardiography was performed according to The American Society of Echocardiography recommendations (8). Resting heart rate was measured using 12-lead electrocardiograms. Prescribed doses of loop diuretics, angiotensin converting enzyme inhibitors (ACEi), angiotensin receptor blockers (ARB) and β-adrenoceptor antagonists (β-blockers) were collected at study recruitment. Total daily doses of β-blockers, ACEi (or ARB if used instead of ACEi) and loop diuretic were expressed relative to the maximal licensed dose of bisoprolol, ramipril and furosemide, respectively, as previously published (4). Receipt of cardiac resynchronisation therapy (CRT) or implantable cardioverter-defibrillator (ICD) was assessed during the six-month period after recruitment.

All patients were registered with the UK Office of Population Censuses and Surveys, which provided details of time of death, with a final censorship date of 8^th^ November 2018; maximum follow-up was for 11 years. Actuarial survival predictions were derived from the United Kingdom National Life Tables (UK-NLT), an official survival estimation measure produced by the UK government (9). The UK-NLT provide annual death rates by sex and age for overlapping three-year periods, which we assigned the value to the middle of the range: for example, the death rate for 2011-2013 is used with patients recruited in 2012. This provides the baseline survival for members of the public with this age and sex, which we used as a reference control population.

### Statistics

Patient characteristics are reported using the mean and standard deviation for continuous variables, with categorical variables summarised using the count of each class and the percentage of the dataset it represents. Median survival rates and Kaplan-Meier curves describing the absolute survival, stratified by sex, were produced using the survival package in R (https://CRAN.R-project.org/package=survival). Relative survival results were produced using the relsurv package within R (https://www.jstatsoft.org/article/view/v087i08). In particular, we use relative survival tables to investigate the excess loss of life associated with heart failure, both on the entire cohort, stratified by sex, and on the subset of participants according to the number of comorbidities. Wald confidence intervals are used for mortality rate, whilst 500 bootstrap samples are used to produce confidence intervals for years of life lost, with a t-test to compare the means between sexes. To investigate the additional impact of comorbidities, an additive relative survival model was produced using the relsurv package within R. The presence of four major comorbidities (COPD, diabetes, ischaemic aetiology, CKD grade ≥4) were used as independent variables. Excess hazard ratios and Wald confidence intervals are reported, fit using the maximum likelihood principle. A sensitivity analysis was performed to determine whether the excess hazard due to heart failure is best modelled as constant or time-varying: we compare a constant term with piecewise constant on one-year intervals and a continuous variant fit using the expectation maximization approach.

## Results

As described in **Table 1**, the study cohort had a mean age of 69.6 years and 73% were male. The aetiology of heart failure was ischaemic heart disease in 59% of cases, mean left ventricular ejection fraction was 32%, and 31% of people had moderate to severe dyspnoea (NYHA classification 3 or 4). Major comorbidity was common, with diabetes being present in 28%, COPD in 16%, and CKD grade ≥4 in 18%. After a maximum follow-up period of 11 years (median 5-years), 999 (55.7%) deaths occurred. As illustrated in **Figure 1A**, median survival was 6.6 years (95% confidence interval 6.3 to 7 years). However, this illustrates a composite of the excess risk of death in this cohort *plus* the background risk in the general population, which is likely to be substantial in the context of their advanced age. To address this, we constructed relative survival models that define the expected loss of survival in the background population, and therefore the excess risk in the study cohort (**Figure 1B**). After 10 years, the expected background population mortality rate is 28.6% (95% confidence interval [CI] 27.8-29.4%); in addition to this, our study cohort experienced an excess risk of 37% (95% CI 33.6-40.5%). Expressed as years of life lost over 10 years of follow-up, the expected loss accounts for 1.6 (95% CI 1.54-1.72) years, whilst the excess risk accounts for a further 2.2 (95% CI 1.99-2.41) years, resulting in a cumulative loss of 3.8 (95% CI 3.66-4.0) years. Therefore, our study cohort lost 2.4-fold (95% CI 2.2-2.5) more life than expected.

**Table 1:** Participant characteristics.

|  | **Total cohort**<br>**(n=1794)** | **Men**<br>**(n=1311)** | **Women**<br>**(n=483)** | **p value** |
| --- | --- | --- | --- | --- |
| Age (years) | 69.6 (12.5) | 69.3 (12.1) | 70.4 (13.5) | 0.1 |
| Male sex (n [%]) | 1311 (73.1) | N/A | N/A | N/A |
| Ischaemic aetiology (n [%]) | 1064 (59.3) | 835 (63.7) | 229 (47.4) | <0.001 |
| Diabetes (n [%]) | 504 (28.1) | 384 (29.3) | 120 (24.8) | 0.06 |
| COPD (n [%]) | 283 (15.8) | 195 (14.9) | 88 (18.2) | 0.09 |
| CKD 4 or above (n [%]) | 141 (7.9) | 86 (6.6) | 55 (11.4) | 0.001 |
| NYHA class 3/4 (n [%]) | 551 (30.7) | 386 (29.5) | 165 (34.2) | 0.06 |
| LV ejection fraction (%) | 32 (9.5) | 31.7 (9.5) | 32.6 (9.5) | 0.08 |
| Betablocker use (n [%]) | 1516 (84.7) | 1117 (85.5) | 399 (82.6) | 0.14 |
| Bisoprolol equivalent dose (mg/day) | 3.9 (3.4) | 4 (3.4) | 3.5 (3.3) | 0.01 |
| ACEi or ARB use (n [%]) | 1618 (90.4) | 1195 (91.4) | 423 (87.6) | 0.014 |
| Ramipril equivalent dose (mg/day) | 4.9 (3.5) | 5.1 (3.6) | 4.3 (3.4) | <0.001 |
| MRA use (n [%]) | 684 (38.2) | 507 (38.8) | 177 (38.8) | 0.41 |
| Furosemide equivalent dose (mg/day) | 51 (50) | 52 (52) | 49 (43) | 0.18 |
| CRT (n [%]) | 452 (25.2) | 353 (26.9) | 99 (20.5) | 0.005 |
| ICD (n [%]) | 209 (11.6) | 184 (14) | 25 (5.2) | <0.001 |

Continuous data displayed as mean (standard deviation) and categorical data as number (%). COPD – chronic obstructive pulmonary disease; CKD – chronic kidney disease; NYHA – New York heart association; ACEi – angiotensin converting enzyme inhibitor; ARB – angiotensin receptor blocker; MRA – mineralocorticoid receptor antagonist; CRT – cardiac resynchronisation therapy; ICD – implantable cardioverter-defibrillator.

**Figure 1:**
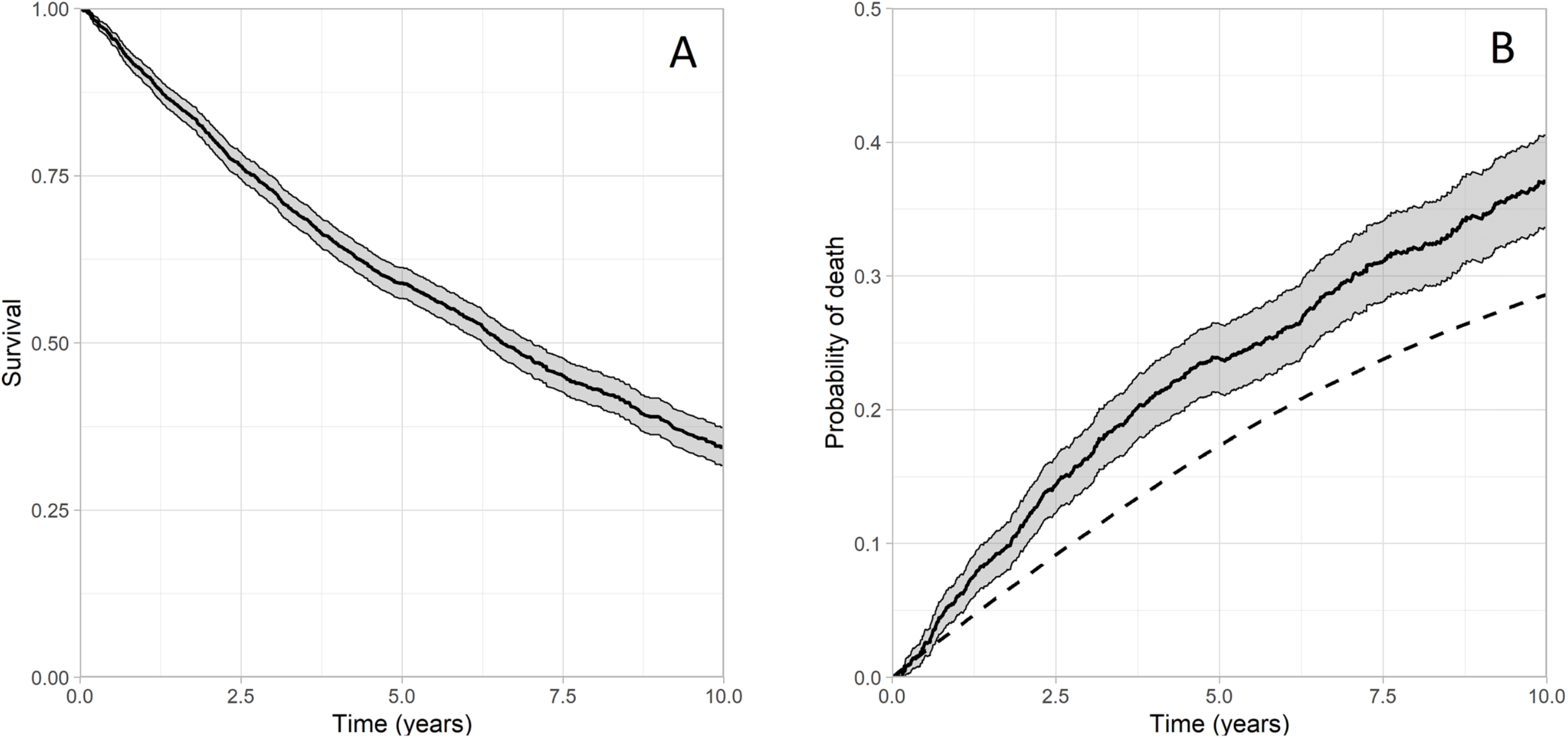
Absolute and relative survival of the study cohort. Legend: **A**) Kaplan-Meier curve illustrating cohort survival (solid line with grey 95% confidence interval); **B**) Relative survival curve illustrating excess mortality in cohort (solid line with grey 95% confidence interval) and projected mortality in an age-sex matched reference control population (dashed line).

Next we explored the impact of male sex, given its established role as an adverse prognostic factor. Relative survival curves for our cohort stratified by sex are given in **Figure 2**. When the expected background and excess mortality were defined with relative survival tables, men and women exhibited similar 10-year background mortality rates (27.9% [26.9-28.9%] versus 30.5% [29-32.1%]). However, excess 10-year mortality rates were higher in men than women (40.3% [36.3-44.2%] versus 28% [21-35.1]). Over 10-years of follow-up, the background loss of life was 1.6 years in both men and women, but the excess risk was 2.4 (95% CI 2.2-2.7) years in men versus 1.6 (95% CI 1.2-2.0) years in women, resulting in an average cumulative loss of 4 and 3.2 years, respectively. Therefore, men and women lost 2.5-fold (95% CI 2.3-2.7) and 2-fold (95% CI 1.7-2.3) more life than expected, respectively, suggesting male sex is associated with a higher-risk heart failure phenotype (p<0.001).

**Figure 2:**
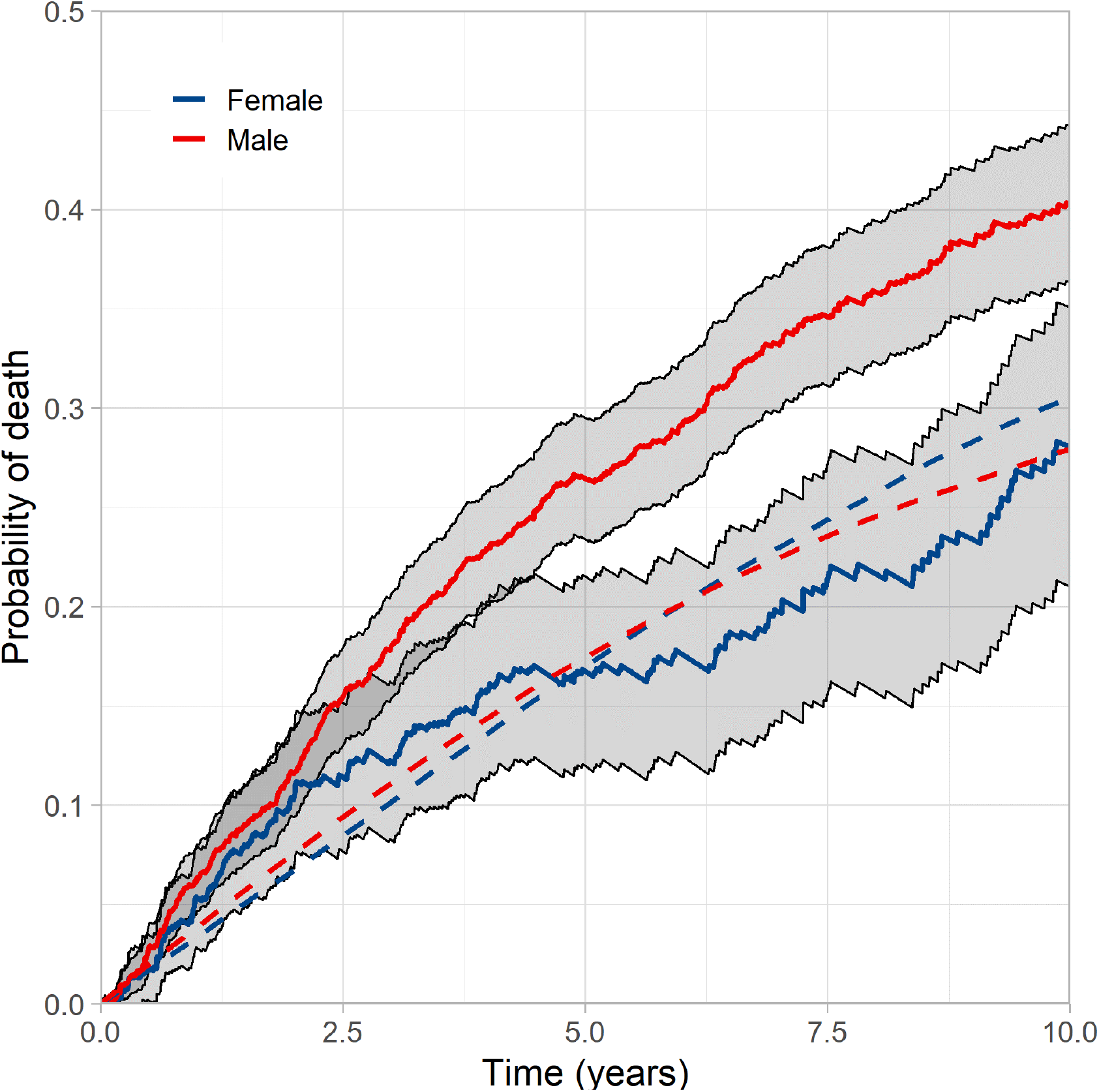
Relative survival stratified by gender. Legend: Relative survival curves illustrating excess mortality in men and women (red and blue solid lines, respectively, with grey 95% confidence intervals) and projected mortality in an age matched reference control population (red and blue dashed lines, respectively).

Given the differing comorbidity profile of men and women (**Table 1**), we next explored how they might contribute to the differential loss of expected life in these groups. As illustrated in **Figure 3**, men and women with increasing numbers of comorbidities experienced substantially greater loss of life expectancy. Indeed, in patients with 3 or more comorbidities, men lost an excess of 4.6 years (95% CI 3.1-5.5), whilst women lost an excess of 3.1 years (95% CI 1.9-4). Importantly, in patients without major comorbidity, men still experienced excess loss of life, whilst women experienced less and were non-significantly different from the reference population (1 year [95% CI 0.6 to 1.5] versus 0.4 years [95% CI -0.3 to 1]; p<0.001). To explore the contribution of specific comorbidities to loss of expected life, a multivariate Cox regression analysis was performed and the excess hazard ratios (EHRs) are presented in **Table 2**. All of these were associated with loss of expected life, but with substantial heterogeneity in their effect size. Notably, whilst statistically significant, the baseline excess hazard was small and approximately constant for the duration of the study; this implies that the excess risk associated with heart failure *per se* remained broadly constant. Moreover, sensitivity analyses using various approaches to allow time-variance in the baseline excess hazard did not reveal differences in the EHRs of the main comorbidities.

**Table 2:** Multivariate survival analysis.

|  | **EHR** | **95% CI of EHR** |  | **p-value** |
| --- | --- | --- | --- | --- |
|  |  | **Low** | **High** |  |
| Diabetes | 1.78 | 1.44 | 2.20 | <0.001 |
| COPD | 2.58 | 2.06 | 3.24 | <0.001 |
| Ischaemic aetiology | 1.42 | 1.13 | 1.78 | 0.004 |
| CKD 4 or above | 2.77 | 2.10 | 3.66 | <0.001 |
| Baseline | 0.053 | 0.048 | 0.060 | <0.001 |

Excess hazard ratios describe risk of reduced life expectancy relative to actuarial projections. CI – confidence interval; EHR – excess hazard ratio; COPD – chronic obstructive pulmonary disease; CKD – chronic kidney disease.

**Figure 3:**
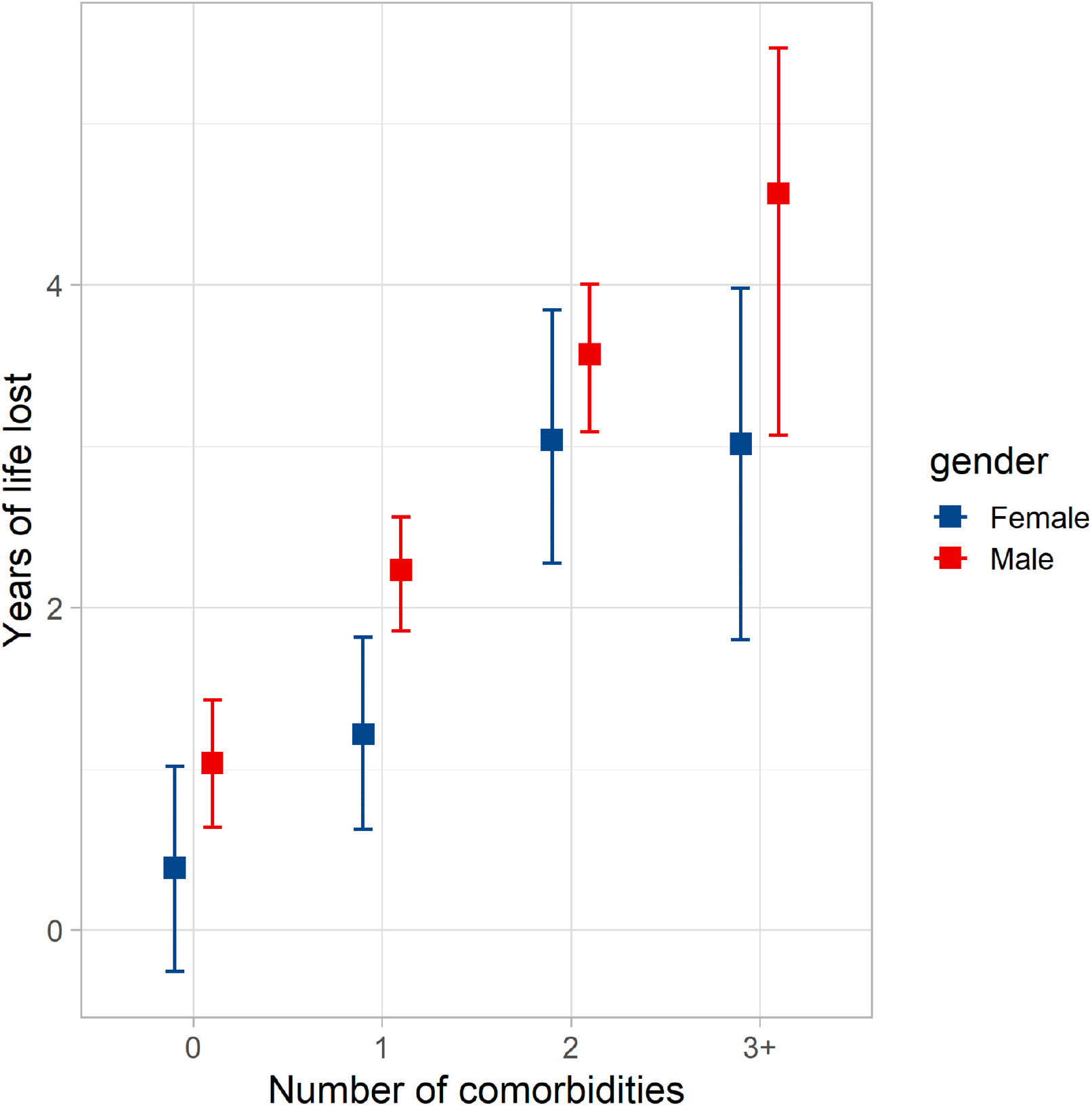
Loss of expected life according to sex and number of co-morbidities. Legend: Loss of expected life over 10-years of follow-up, with 95% confidence interval, in men (red) and women (blue) according to number of comorbidities (from ischaemic heart disease, chronic obstructive lung disease, diabetes, and chronic kidney disease stage 4 or above).

## Discussion

By considering survival relative to actuarial estimates of life expectancy, we have shown that heart failure is associated with a 2.4-fold greater loss of time alive than observed in the age-sex matched general population over ten years. Notably, male sex and accrual of major comorbidities are associated with larger loss of life, whilst women without major comorbidity have life expectancy compatible with actuarial projections. This approach to defining survival may provide useful perspective for clinicians considering the magnitude of risk posed by heart failure in the context of an aging and increasingly multimorbid population. This context may be particularly useful when communicating risk to people with heart failure, who often struggle to estimate their own prognosis.

### Estimating prognosis

Validated tools, such as the Seattle Heart Failure Model (SHFM) and the Meta-analysis Global Group in Chronic Heart Failure (MAGGIC) score (10,11), are already available to estimate the prognosis of people with heart failure in terms of absolute lifespan. Whilst valuable, it is important to ask whether this approach tells patients and clinicians what they want to know. By overlooking the inevitability of death in similar people without disease, such prognostic estimates may be misinterpreted, resulting in poorly informed decision making. The challenges of prognostication in people with heart failure are illustrated by the discordance between model- and patient-estimated absolute life expectancy (6). By considering survival relative to actuarially predicted life expectancy, we hope that our approach will provide essential context to aid the challenging process of communicating prognosis. This may take the form of ‘ballpark’ estimates of excess loss of life for groups of similar people, or by developing an individualised prognostication tool, such as the SHFM. Further research is needed to address the validity, acceptability, and added value of this approach, but we think that it has the potential to improve prognostication in clinical practice.

### Multimorbidity as risk marker and therapeutic target

Recent research describing all people with heart failure in a representative cohort of 4 million UK residents found that multimorbidity is becoming increasingly common (2). Whilst we focussed on just four major comorbidities, 26% of our cohort were not multimorbid (i.e. heart failure with at least one comorbidity), and 31% had 2 or more of these comorbidities. Strikingly, people with 3 or more comorbidities experienced approximately 5-fold greater excess loss of life than people with no comorbidity (**Figure 3**). This suggests that the accumulation of comorbidity is an important part of the adverse prognostic impact of heart failure. Optimal medical therapy is associated with substantial reductions in heart failure morbidity and lifespan extension in clinical trial participants (12), yet clinical trials often exclude multimorbid people. These data highlight the need to design clinical trials specifically recruiting people with heart failure and multimorbidity, possibly applying complex interventions that target more than just the heart failure syndrome.

### Heart failure in men and women

Poorer survival of men has been observed in many studies of heart failure, and is accounted for in the SHFM and MAGGIC prognostic models (10,11). Whilst this could to some extent be attributed to differences in comorbidity, such as ischaemic heart disease, our observations from people with heart failure and no major comorbidity still show clear differences in the outcomes of men and women. Notably, the survival of women without major comorbidity overlapped with that of the matched general population (**Figure 3**). The mechanisms of this sexual dimorphism remain debated (13,14), but it is clear that clinical trials and guidelines should carefully consider the differences between men and women with heart failure.

### Limitations

Although our work has key strengths, it is important to acknowledge limitations that should be addressed by ongoing research. First, we have deliberately chosen not to derive an individualised risk assessment tool, as the aim of this paper is to describe survival relative to life expectancy in populations with heart failure. However, our methods could easily be used to extend the data provided by individualised prognostic models, such as SHFM and MAGGIC (10,11). It will also be important to understand whether health care professionals and patients find survival estimates relative to actuarial life expectancy more useful than absolute survival estimates. Next, our data should not be generalised to other populations (e.g. heart failure with preserved ejection fraction), but our methods could easily be applied to published datasets. It is also important that our 11-year follow-up period represents a modest proportion of predicted life expectancy in our youngest participants, so caution should be applied in extrapolating our data to the youngest people with heart failure. Finally, it is important to note that our expected survival data are derived from the UK general population which will include some people with heart failure; therefore, loss of expected survival is in relation to the age-sex matched general population, not an age-sex matched heart failure free population.

### Conclusions

By framing survival in the context of actuarial predictions, we have shown that people with heart failure with reduced left ventricular ejection fraction lose 2.4-fold more of life than expected. However, most of this loss of life expectancy is accounted for by people with comorbidity, particularly in women. Our work provides a different framework for clinicians and people with heart failure to consider prognosis and should prompt more focus on the issue of heart failure associated with complex multimorbidity.

## Data Availability

The datasets generated and/or analysed during the current study are not publicly available due to inclusion of potentially identifying postal codes, but are available from the corresponding author on reasonable request

## Abbreviations

ACEi: Angiotensin converting enzyme inhibitors
ARB: Angiotensin receptor blockers
β-blockers: β-adrenoceptor antagonists
CHF: Chronic heart failure
CRT: Cardiac resynchronisation therapy
ICD: Implantable cardioverter-defibrillator
CKD: Chronic kidney disease
COPD: Chronic obstructive pulmonary disease
eGFR: Estimated glomerular filtration rate
EHRs: Excess hazard ratios
LVEF: Left ventricular ejection fraction
MAGGIC: Meta-analysis Global Group in Chronic Heart Failure
NYHA: New York Heart Association
SHFM: Seattle Heart Failure Model
UK: United Kingdom
UK-NLT: United Kingdom National Life Tables

## Declarations

### Ethics approval and consent to participate

The Leeds West Research Ethics Committee provided ethical approval (07/Q1205/17), and all patients provided written informed consent to participate

### Consent for publication

Not applicable

## Authors’ contributions

MD collected data, analyzed data and drafted the manuscript. SDR collected data, analyzed data and drafted the manuscript. AMNW collected data and drafted the manuscript. TS collected data and critically revised the manuscript. JG collected data and critically revised the manuscript. MFP collected data and critically revised the manuscript. JL collected data and critically revised the manuscript. SS collected data and critically revised the manuscript. AK collected data and critically revised the manuscript. MG collected data and critically revised the manuscript. ADS collected data and critically revised the manuscript. VKG collected data and critically revised the manuscript. RJS collected data and critically revised the manuscript. KKW collected data and critically revised the manuscript. MTK collected data and critically revised the manuscript. RMC collected data, analyzed data, and drafted the manuscript.

## Acknowledgements

We are grateful for support from the NIHR funded clinical research facility at Leeds Teaching Hospitals NHS Trust.

